# Connecting intermediate phenotypes to disease using multi-omics in heart failure

**DOI:** 10.1101/2024.08.06.24311572

**Authors:** Anni Moore, Rasika Venkatesh, Michael G. Levin, Scott M. Damrauer, Nosheen Reza, Thomas P. Cappola, Marylyn D. Ritchie

## Abstract

Heart failure (HF) is one of the most common, complex, heterogeneous diseases in the world, with over 1-3% of the global population living with the condition. Progression of HF can be tracked via MRI measures of structural and functional changes to the heart, namely left ventricle (LV), including ejection fraction, mass, end-diastolic volume, and LV end-systolic volume. Moreover, while genome-wide association studies (GWAS) have been a useful tool to identify candidate variants involved in HF risk, they lack crucial tissue-specific and mechanistic information which can be gained from incorporating additional data modalities. This study addresses this gap by incorporating transcriptome-wide and proteome-wide association studies (TWAS and PWAS) to gain insights into genetically-regulated changes in gene expression and protein abundance in precursors to HF measured using MRI-derived cardiac measures as well as full-stage all-cause HF. We identified several gene and protein overlaps between LV ejection fraction and end-systolic volume measures. Many of the overlaps identified in MRI-derived measurements through TWAS and PWAS appear to be shared with all-cause HF. We implicate many putative pathways relevant in HF associated with these genes and proteins via gene-set enrichment and protein-protein interaction network approaches. The results of this study (1) highlight the benefit of using multi-omics to better understand genetics and (2) provide novel insights as to how changes in heart structure and function may relate to HF.

## 1. Introduction

### 1.1. Heart failure has a high disease burden

Heart failure (HF) carries one of the highest disease burdens in the world, with 1-3% of the global population estimated to be living with HF. This includes 6.7 million people in the United States (US) alone, and does not include the 33% of the US population at-risk of developing HF^1^. The etiology of HF is heterogeneous and complex, but has ultimately been defined as a clinical syndrome with symptoms and signs caused by structural and functional cardiac abnormalities^2^. Its risk is promoted by increasing age and by the presence of comorbidities such as myocardial infarction, diabetes, hypertension, obesity, arrhythmias, infiltrative and inflammatory disorders, and exposure to drugs or environmental toxins^3–5^. Despite the complexity of HF, it has been demonstrated that risk is impacted by genetic predisposition to some degree^6^. While an exact consensus of heritability for HF has not been reached, some have estimated that the heritability of HF is around 26%^7^.

The overall progression of HF can be broken up into four stages:

Stage A: risk of HF but patients have no symptoms or structural heart changes
Stage B: no symptoms of HF or asymptomatic but patients do have structural heart changes
Stage C: patients experiencing symptoms of HF
Stage D: advanced heart failure requiring specialized interventions^8,9^.

As noted in Stage A and B, HF is often preceded by a phase of undetected progression, highlighting the need for better insight into the changes, such as structural heart changes^9,10^. These changes often appear specifically at the left ventricle (LV), and include decreased left ventricular ejection fraction (LVEF), LV dilation and/or hypertrophy, and valvular disease in which the heart cannot pump as effectively, losing function. LV mass (LVM) has been shown to be an independent predictor of HF, with risk for HF increasing by 1% for every 1% increase in excess LV mass^11^. Likewise, LVEF, which measures LV contractile function (the percentage of blood leaving the heart with each contraction) has been shown to be associated with HF prognosis^12,13^. Abnormal measurements of several of these parameters measuring both structural and functional changes together are reliable markers of cardiovascular risk and eventual HF diagnosis.

To quantify changes in the volume of blood in the heart before and after contraction, we can use LV end-diastolic volume (LVEDV) and LV end-systolic volume (LVESV) respectively. Together these four measures of heart structure and function (LVM, LVEF, LVEDV, and LVESV) can provide an overall characterization of progression towards potential HF and represent an intermediate phenotype or endophenotype. Identifying overlaps in changes seen in both intermediate MRI trait measures and HF could give us a better idea of vital aspects that lead towards full, advanced HF.

### 1.2. Using multi-omics to increase knowledge gained from GWAS

Given the known genetic contribution to HF and the prevalence of patients with the disease, many groups have performed genome-wide association studies (GWAS) to identify genetic variants associated with HF^14–21^. While this approach allows us to gain valuable insights into potential genetic variation that contributes to the disease, it still leaves a crucial gap in connecting how these variants are actually resulting in mechanistic change, and in which specific tissues. This is especially relevant in quantitative phenotypes, where GWAS is insufficient to capture the full heterogeneity measured by the trait. Transcriptome-wide association studies (TWAS) use GWAS summary statistics along with reference gene expression from specific tissues to predict how genetic variants affect gene expression within those tissues. TWAS also provides a boost in overall statistical power, as it is less affected by multiple test corrections due to being a gene-based test of association^22,23^. This information gain increases further with the use of protein-wide association studies (PWAS), which use protein expression measures from tissue as a reference to predict genetic changes in protein abundance, and further cut down the multiple test correction as it is a gene (protein) based test. These methods are also more portable than GWAS; they are less impacted by population structure in datasets as they operate on a gene and protein level^24^.

In this study, we make use of TWAS and PWAS methods to investigate genetic-derived gene and protein changes among cardiovascular related-tissues using the largest published GWAS summary statistics of HF and MRI measures of LV structure and function to date^25–27^. Our goals are: 1) to integrate multi-omics data in the form of reference gene expression and protein expression datasets to identify novel HF and related trait associated genes, 2) to evaluate whether TWAS and PWAS approaches uncover the same association signals or provide novel gene-based associations, and 3) determine whether these genes associated with HF and related traits are part of shared pathways and/or networks between traits. This study is also, to our knowledge, one of the first times that both TWAS *and* PWAS have been performed simultaneously on quantitative traits.

## 2. Methods

### 2.1. Cardiovascular data

2.1.1. *MRI traits*

Of all cardiac chambers, dysfunction of the left ventricle is the most common structural abnormality in HF cases. We chose four measurements taken from the left ventricle derived from MRI imaging with previously published GWAS data to characterize potential associations with HF: LVM indexed to body surface area, LVEDV, LVESV, and LVEF^26,27^. LVM measurements were taken from Khurshid et al, and includes 43,230 samples (91% European ancestry) with MRI imaging and genotype data from the UK Biobank^26^. LVEDV, LVESV, and LVEF association studies were performed on 41,135 samples also from the UK Biobank with MRI imaging and genotype data by Pirruccello et al^27^.

2.1.2. *Heart failure (HF)*

We identified the largest all-cause HF GWAS study to date including 207,346 non-overlapping samples of cases and 2,151,210 controls meta-analyzed from HERMES, the Million Veterans Project (MVP), FinnGen, Mount Sinai BioMe (BIOME), Global Biobank Meta-analysis Initiative (GBMI), eMERGE, Geisinger DiscovEHR, and Penn Medicine BioBank (PMBB)^25^. This included an overall sample of 81.1% European ancestry, 9.7% African American, 6.5% East Asian, and 2.6% Admixed American.

### 2.2. Transcriptome-wide association study (TWAS)

To provide tissue-specific context to GWAS results from the selected MRI traits and HF studies we conducted transcriptome-wide association studies (TWASs) using S-PrediXcan^28^ and multivariate adaptive shrinkage (MASHR) eQTL models from the Genotype-Tissue Expression (GTEx) Project v8, available in PredictDB^29,30^ GTEx eQTLs were derived from a sample group of mostly European ancestry (84.6% European ancestry, 12.9% African American, 1.3% Asian and 1.1% unknown) that closely parallels the composition of the HF multi-ancestry cohort. Using this reference, we imputed genetically regulated gene expression (GReX) in ten tissues known to be relevant in the cardiovascular system and heart failure (aorta, coronary artery, tibial artery, atrial appendage, left ventricle, whole blood, visceral adipose, subcutaneous adipose, liver, and kidney)^31–37^. Associations for each of the intermediate MRI traits from the UK Biobank, as well as multi-ancestry and EUR populations from all-cause HF were calculated independently for each of these ten tissues. Significant genes were determined using a Bonferroni threshold of (p<0.05/# genes tested) per trait. TWAS genes for all traits were then fine-mapped using FOCUS^38^ to eliminate genes significant due to linkage disequilibrium (LD).

### 2.3. Proteome-wide association study (PWAS)

We performed a proteome-wide association study (PWAS) using S-PrediXcan^28^ with the GWAS summary statistics for MRI traits from UK Biobank and for the multi-ancestry and European (EUR) population all-cause HF studies. PWAS identifies genetic associations that may influence complex traits, such as all-cause HF and MRI traits, by regulating protein abundance in tissue^39^. Blood plasma-derived protein quantitative trait loci (cis-pQTLs) from the Atherosclerosis Risk in Communities (ARIC)^40^ study were used to construct the models. This large bi-ethnic study was made up of 9,084 participants, consisting of 7,213 European Americans (EA) and 1,871 African Americans (AA). S-PrediXCan PWAS EA and AA models were identified in PredictDB and were constructed using ARIC consortium data by utilizing PEERS covariates, expression information from eQTL associations, gene and SNP annotations^41,42^. PWAS was conducted on multi-ancestry and EUR studies of all-cause HF, as well as on traits from UK Biobank with the intermediate MRI traits using the EA cohort information, and additionally the AA cohort for the multi-ancestry HF study. The resulting PWAS associations were assessed for statistical significance using a Bonferroni significance threshold (p<0.05/# proteins tested) for each trait.

### 2.4. Network and pathway analyses

#### 2.4.1. Pathway enrichment analysis

Gene set enrichment was performed using EnrichR^43,44^ for the significant results from TWAS and PWAS for each MRI trait and HF phenotype, respectively. Enrichment analysis explored the specific pathways and processes associated with the statistically significant genes and proteins from the TWAS and/or PWAS. Pathway results were annotated with KEGG 2021, Reactome 2022, and Gene Ontology (GO) Biological Process 2023 pathways. The significant pathways were identified as having Fisher’s exact test p-value < 0.05^39,45^.

#### 2.4.2. Network analysis and identification of hub genes and proteins

The statistically significant genes and proteins identified via TWAS and PWAS were used to construct a protein-protein interaction (PPI) network using the online Search Tool for the Retrieval of Interacting Genes (STRING v11)^46^, where the number of interactions present was assessed for significance. Network interactions were thresholded by a minimum confidence score of > 0.4, as calculated by STRING^46^. The networks were then visualized using Cytoscape 3.10.2^47^, and degree centrality analysis was performed using the cytoHubba module to identify and visualize the hub genes and proteins^48,49^.

#### 2.4.3. Classification of of sub-clusters

Additionally, the Molecular Complex Detection (MCODE)^50^ module in Cytoscape was used to screen modules of the larger PPI networks and construct clusters by identifying densely-connected regions of the network^51^. The networks were thresholded to have an MCODE degree cutoff of 3, node density cutoff of 0.1, node score cutoff of 0.2, number of nodes > 3^48,52^. Gene set enrichment analysis using KEGG 2021^53–55^, Reactome 2022^56,57^, and Gene Ontology (GO) Biological Process 2023^58,59^ of each cluster was then conducted using Metascape^60^, using the default parameters of minimum overlap of 3, p-value cutoff of 0.01, and minimum enrichment score of 1.5.

## 3. Results

### 3.1. TWAS and PWAS Association Analyses

#### 3.1.1 MRI trait gene and protein associations

After fine-mapping, 35 unique genes within ten tissues and three proteins from blood plasma (SPON1, C2, PACAP) were significant for LVEF based on a Bonferroni threshold (TWAS:p<3.814E-07, PWAS:3.75E-05) (**Figure 2A**). 16 of these genes were significant in three or more tissues, and one gene, *SPON1*, replicated in both TWAS and PWAS for LVEF. 35 genes and one protein (THBS4) appeared significantly associated with LVM measures (TWAS:p<3.842E-07, PWAS:3.79E-05). Five of these significant genes (*FKBP7*, *WNT3*, *HSPQ4*, *PSMC3*, and *PRKRA*) appeared in three or more tissues tested. Finally, amongst the ten tissues tested, 33 genes and three proteins (ENG, QPCTL, SPON1) were significant for LVEDV (TWAS:p<3.815E-07, PWAS:3.79E-05)and 48 genes along with four proteins (RAB5A, SRL, PACAP, SPON1) for LVESV (TWAS:p<3.815E-07), protein:3.79E-05). *SPON1* was also significantly associated with LVESV for both TWAS and PWAS. Figures for LVM, LVEDV, LVESV are available in **Supplemental Figure 1A-C**. The full significant results of the TWAS and PWAS for MRI traits are available in **Supplemental Table 1 and 2**.

**Figure 1.**
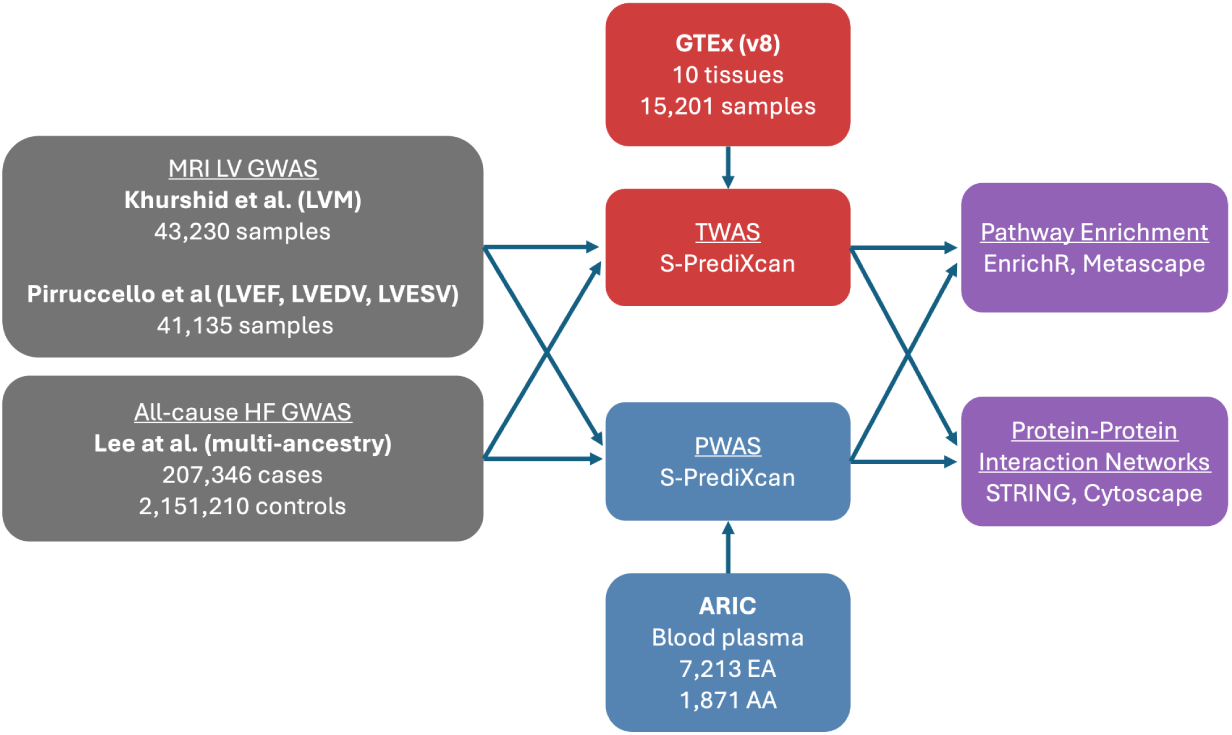
Overview of study analysis to identify genes, proteins, and related interactions between MRI-derived cardiac intermediate traits and heart failure GWAS. (LVM: Left ventricular mass, LVEF: Left ventricular ejection fraction, LVEDV: Left ventricular end-diastolic volume, LVESV: Left ventricular end-systolic volume, EA: European American, AA: African American)

**Figure 2.**
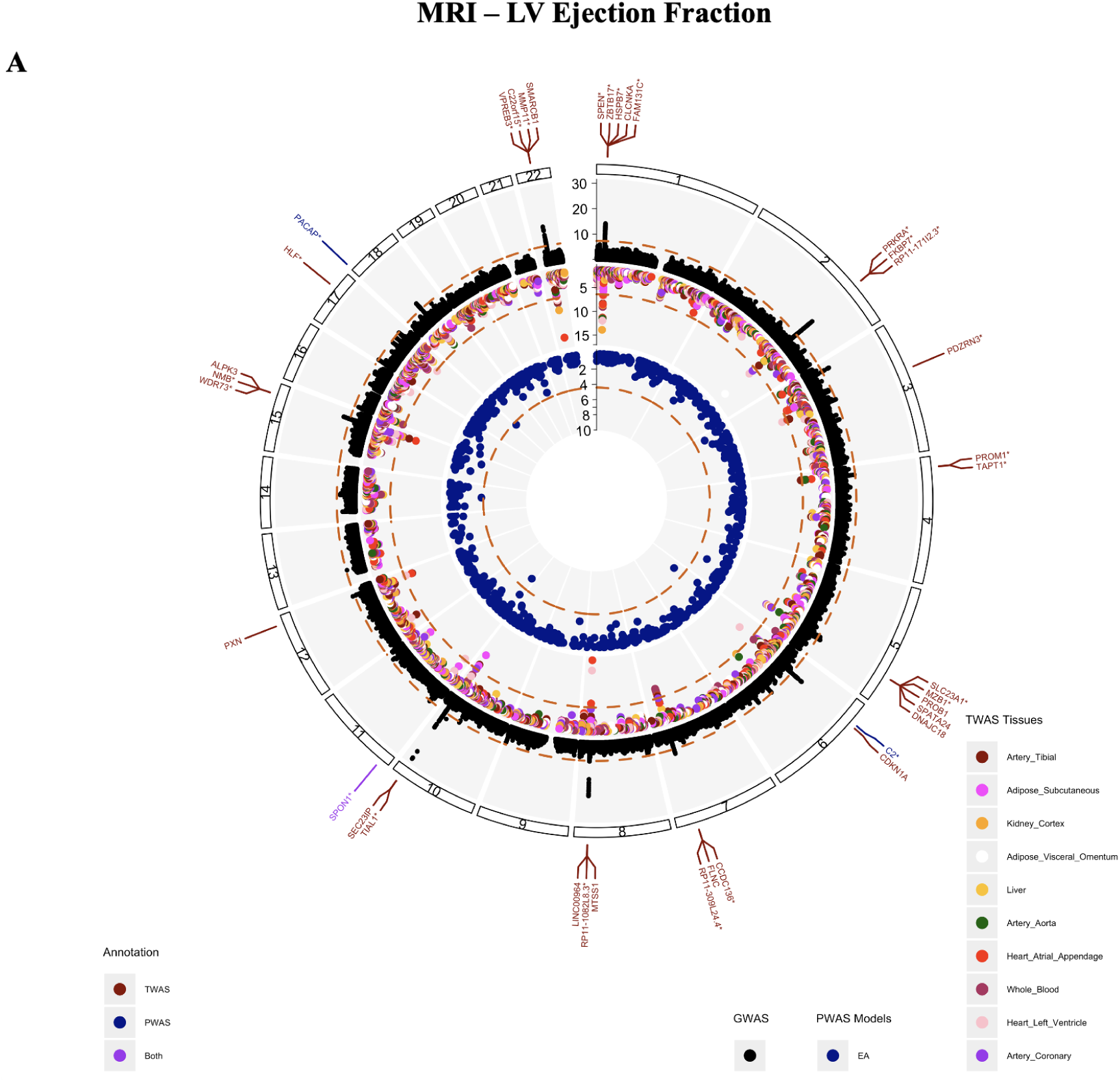
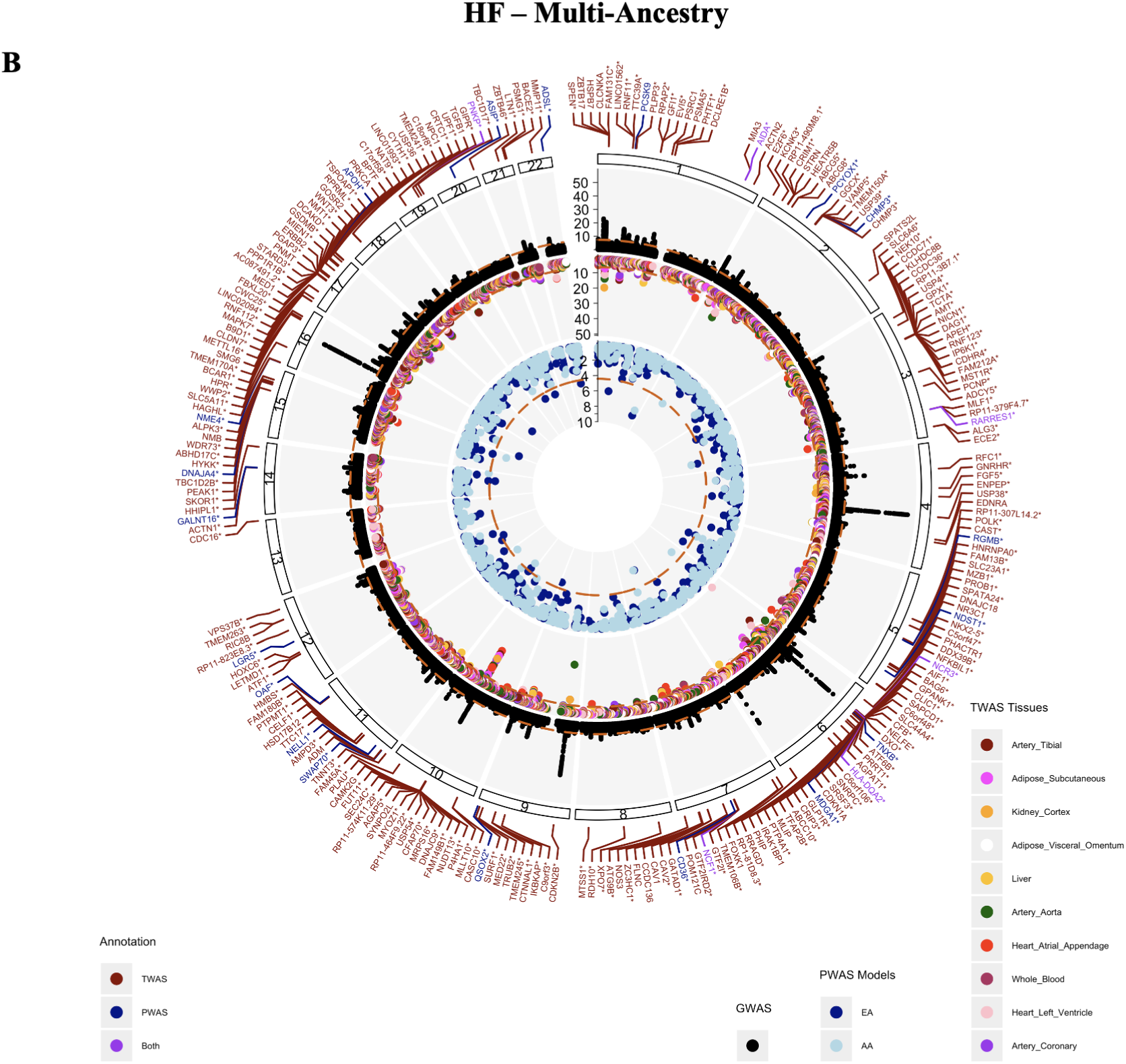
Circos plots for A) MRI LV Ejection Fraction and B) HF - Multi-ancestry representing identified associations through GWAS (black), TWAS (red), and PWAS (blue). The outermost track of annotations depicts genes and proteins identified through TWAS (red) and PWAS (blue), or both analyses (purple), with the asterisk denoting novel results not previously reported from the source GWAS, GWAS Catalog, or NCBI.

Between MRI traits, several genes appeared significant between measures. Genes *FKBP7*, *PRKRA*, and *RP11-171I2.3* were associated with all four MRI-based traits in at least one tissue. 15 genes overlapped between LVEDV and LVESV, four genes were shared between LVEDV and LVM, four between LVEDV and LVM, five genes between LVESV and LMV, and finally 30 genes between LVESV and LVEF. Amongst protein results, SPON1 was the only protein shared between traits and was significantly associated with LVEF, LVEDV, and LVESV.

#### 3.1.2 HF gene and protein associations

231 unique genes and 29 proteins significantly associated with HF in a multi-ancestry population (TWAS:p<3.806E-07, PWAS:p<3.79E-05) (**Figure 2B**). Six genes replicated across genes and proteins, including *RARRES1*, *NCF1*, *AIDA*, *HLA-DQA2*, *PNKP*, and *NCR3*. 185 of the total 231 associating genes were significant in at least one vascular tissue (heart atrial appendage, heart left ventricle, tibial artery, coronary artery, whole blood). 90 genes were significant in at least one vascular tissue and one peripheral tissue (liver, kidney, subcutaneous adipose, visceral adipose). Of the ten tissues tested, heart atrial appendage and heart left ventricle tissues had the largest number of genes significantly associating with HF. Genes *CRIP3* and *USP54* were significant in all ten tissues tested. Similar associations with HF were noted in the European population **(Supplemental Figure 1D).**

### 3.2. Network and Pathway analyses

#### 3.2.1. MRI trait gene-set enrichment

In order to identify the known biologically relevant pathways associated with the statistically significant TWAS and PWAS genes for each phenotype, gene-set enrichment analysis was performed using EnrichR for Reactome 2022, KEGG 2021, and Gene ontology (GO) 2023 pathways. Significant pathways were identified at a p-value < 0.05; the full set of significant pathways for each phenotype are available in **Supplemental Table 3**. For LVEF, the most significant pathway by p-value was positive regulation of actin filament bundle assembly (p = 4.23E-03), made up of genes *PXN* and *MTSS1* **(Figure 3A)**. Several pathways involved in kidney development function were also identified to be significant, such as renal cell filtration differentiation (p-value = 9.47E-03), and nephron tubule development (p = 9.47E-03).

**Figure 3.**
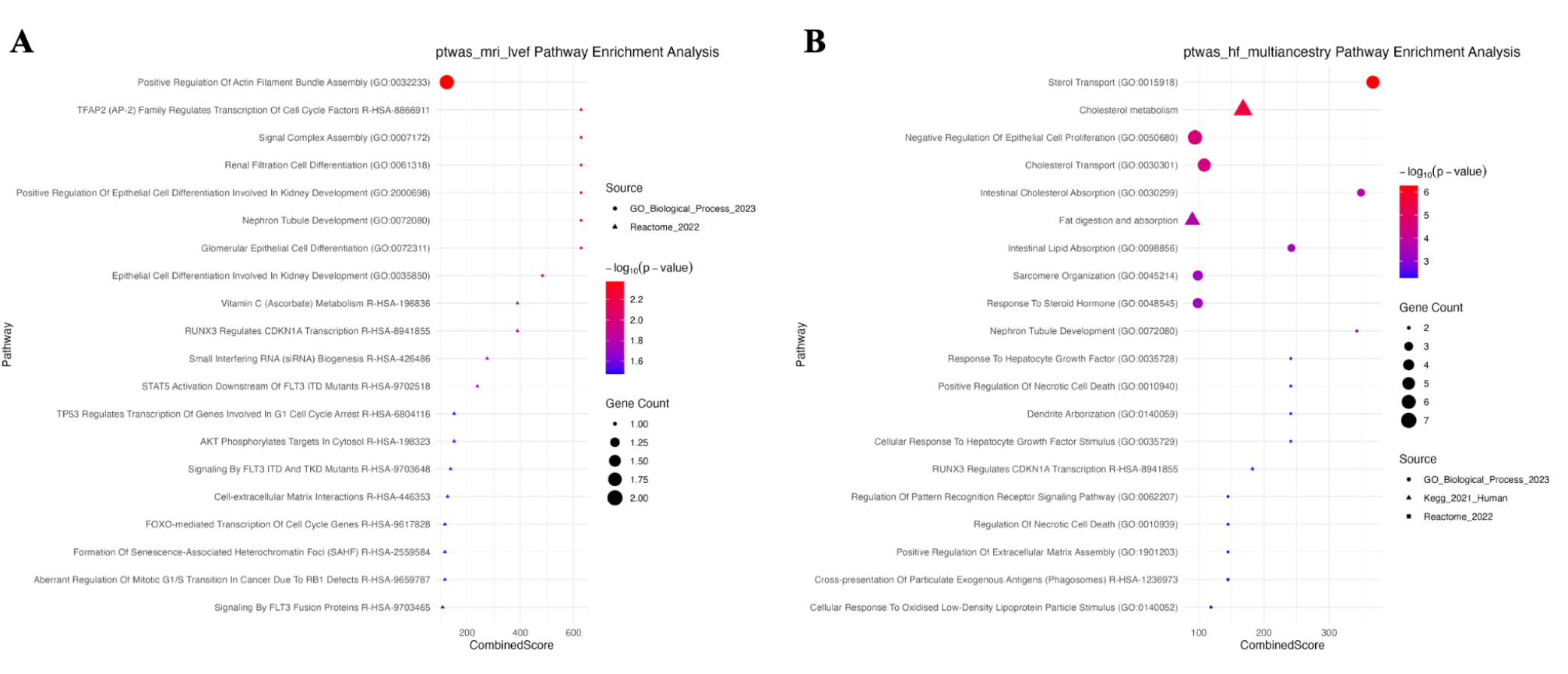
Gene-set enrichment results of TWAS and PWAS significant hits for A) MRI LVEF and B) multi-ancestry HF phenotypes.

LVM **(Supplemental Figure 2A)** was enriched for a variety of cell proliferation and differentiation pathways such as positive regulation of endothelial cell proliferation (p = 1.11E-03) and negative regulation of muscle cell differentiation (p = 4.54E-03), identifying the gene *IGF2* and protein THBS4 as important. Similarly developmentally important cardiovascular pathways were identified for the LVEDV **(Supplemental Figure 2B)**, including embryonic hemopoiesis (p = 2.43E-04), megakaryocyte differentiation (p = 4.211E-04), and cardiac atrium morphogenesis (p = 4.73E-04). The most significant pathway for LVESV **(Supplemental Figure 2C)** was modulation by host of symbiont process (p = 5.34E-03), in addition to developmental pathways - glomerular epithelial cell differentiation (p = 1.29E-02), and renal filtration cell differentiation (p = 1.29E-02).

#### 3.2.2. HF gene-set enrichment

A variety of relevant gene-sets were found to have overrepresented pathways previously identified as important in all-cause HF^61^. For the HF multi-ancestry cohort **(Figure 3B)**, the most significant pathways include sterol transport (p = 5.34E-07) and cholesterol metabolism (p = 2.84E-06), which are known to be impacted in a variety of cardiovascular disease states, including heart failure^62,63^. Similar pathways were enriched in the EUR population for HF **(Supplemental Figure 2D)**. Genes implicated in these pathways include *ABCG8, STARD3, ABCG5, NPC1, CAV1, APOH, PCSK9,* and *CD36*.

#### 3.2.3. PPI network analysis of MRI trait genes and proteins

To evaluate the association of candidate genes and proteins identified by TWAS and PWAS, PPI networks were constructed for each MRI trait phenotype using the STRING database. The PPI network for LVEF contained 32 nodes, 16 of which were connected, and 14 edges at a confidence threshold of > 0.4, with a PPI enrichment p-value = 5.24E-07, indicating that there were significantly more interactions observed than expected by random chance. The hub nodes identified via the cytohubba plugin by degree centrality were FLNC, ALPK3, SPATA24, and HSPB7 (**Figure 4A**). MCODE identified 1 cluster in the network, with nodes FLNC, HSPB7, and ALPK3, at a score of 1.5, as computed by multiplying node density by the number of members.

**Figure 4.**
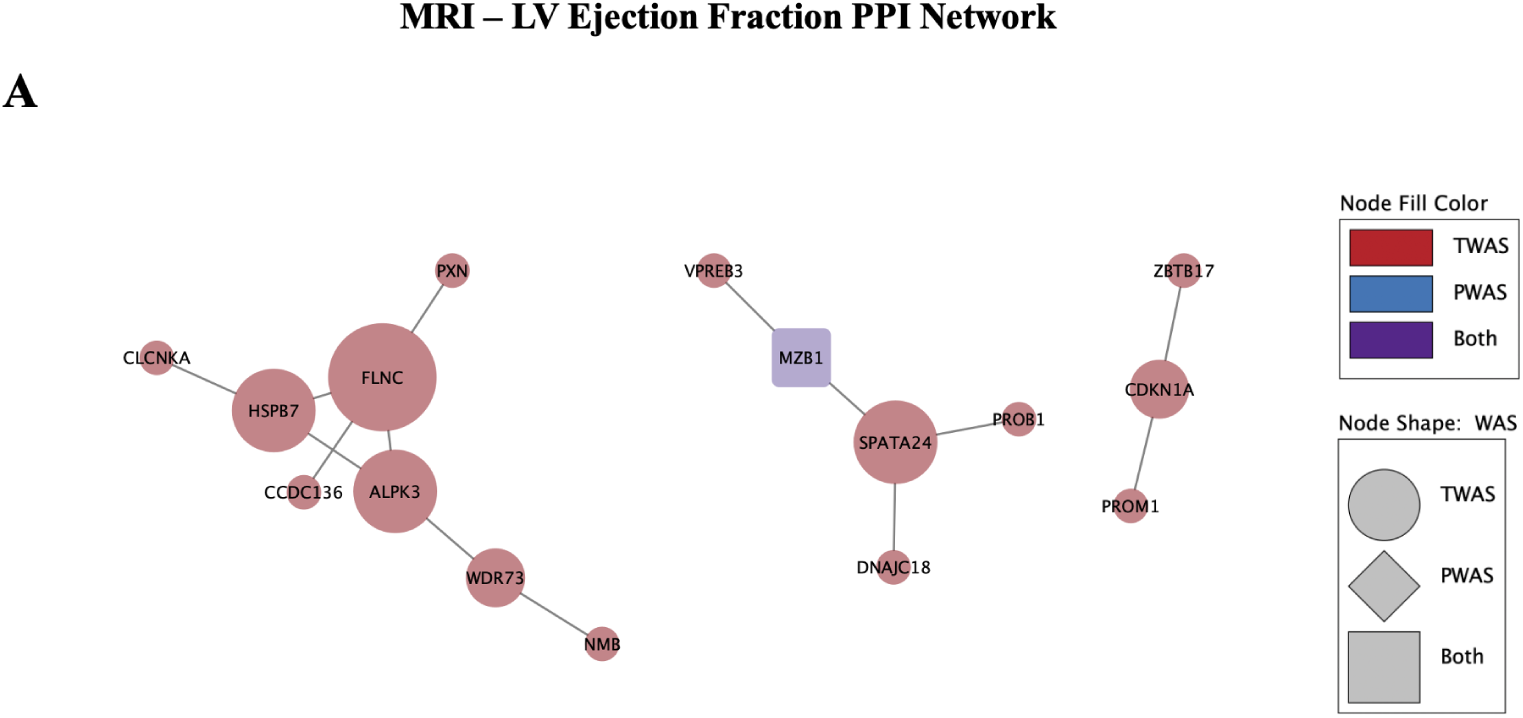
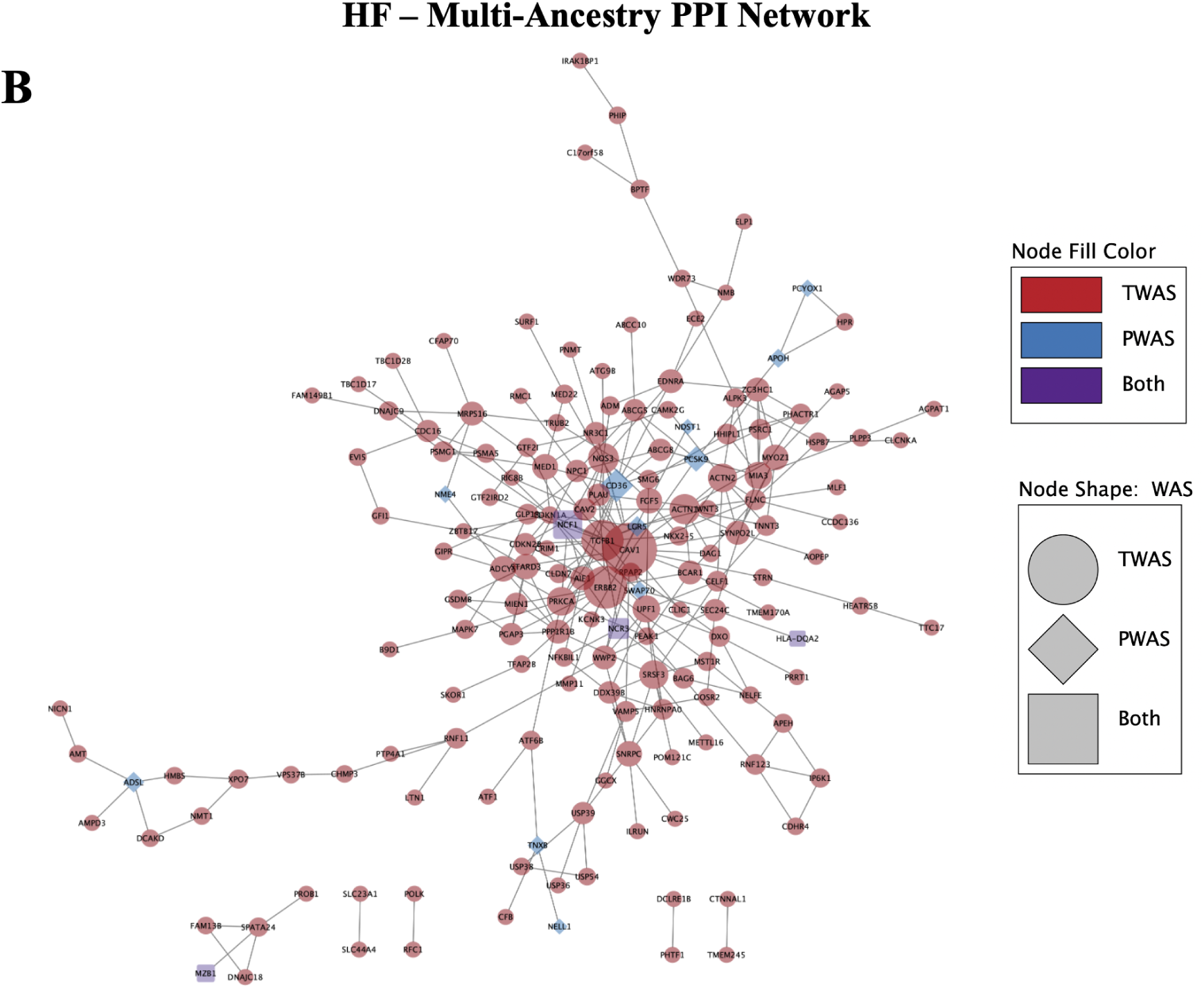
PPI networks constructed using TWAS and PWAS significant hits for A) MRI LVEF and B) multi-ancestry HF phenotypes. Size of nodes denotes degree centrality, with largest nodes identified as hub nodes.

The network for LVM **(Supplemental Figure 3A)** contained 10 nodes (3 connected) and 2 edges, all corresponding to hits from TWAS. The network had a PPI enrichment score of p = 0.303. The hub node identified was FKBP7, a protein which functions as a molecular chaperone to accelerate protein folding. MCODE did not identify clusters in this network.

The network for LVESV **(Supplemental Figure 3B)** consisted of 45 nodes (19 connected) and 16 edges, with a PPI enrichment p-value = 3.14E-06. Similarly to LVEF, the genes FLNC, ALPK3, HSPB7, and BHMG1 were identified as hub nodes. MCODE once again identified 1 cluster in the network, with nodes FLNC, HSPB7, and ALPK3, at a score of 3.

The LVEDV network **(Supplemental Figure 3C)** contained 34 nodes (13 connected) and 11 edges. Cytohubba identified BHMG1 as a hub node. MCODE did not identify clusters in this network. Metascape pathway enrichment of clusters for the MRI phenotypes did not yield additional enriched terms; the full cluster and pathway enrichment results are available in **Supplemental Table 4**.

#### 3.2.4. PPI network analysis of HF genes and proteins

The PPI network for the HF multi-ancestry cohort consisted of 234 nodes and 294 edges at the medium confidence score > 0.4 in STRING, with an average local clustering coefficient of 0.377 (**Figure 4B**). STRING found that the network had a PPI enrichment p-value = 1.0E-16, suggesting that the network had significantly more edges than expected by random chance. In Cytoscape, the hub nodes identified by degree centrality were CAV1, ERBB2, TGFB1, and CD36. Additionally, 5 non-overlapping clusters were identified in MCODE, with a max cluster score of 4.889 and minimum score of 3.0, where a higher score denotes a greater number of nodes in the cluster. Metascape was used to evaluate pathways for the genes present in each cluster, identifying pathways in sarcomere organization (LOG_10_P=-9.09), Hemostasis (LOG_10_P=-7.84), VEGFA-VEGFR2 pathway (LOG_10_P=-7.65), and heart development (LOG_10_P=-3.14), among several others. The PPI network for HF in the EUR population **(Supplemental Figure 3D)** yielded similar findings.

## 4. Discussion

In this study we performed the largest TWAS at the gene level and first-ever protein imputation for all-cause HF. This is also one of the first studies to include intermediate cardiac imaging traits in relation to HF. To contextualize our findings from the TWAS and PWAS, we used associated genes and proteins to identify enriched gene sets and construct interaction networks. Overall we demonstrated (1) an increase in information gain using TWAS and PWAS in addition to GWAS in understanding what kinds of pathways are impacted at the intermediate phenotype and full-stage HF phenotypes, (2) the value in using quantitative intermediate measures for interpreting the changes that occur during disease progression, and (3) how these intermediate measures may relate to changes seen in full-stage HF.

### 4.1. Information gain from multi-omics approaches

Previous GWAS for several MRI-derived cardiac quantitative traits and all-cause HF GWAS have been able to identify a wide range of signals, promoting a broad understanding of genetic factors underlying HF; however, these variant results are unable to give actual clues toward their effects on mechanism as it relates to the traits of interest. Little has been done on the gene level for quantitative cardiac measures as used here. Genes deemed to be significant from GWAS are usually only identified based on their proximity to significant variants, but we know that proximity actually is not always a good indicator of actual effect on expression and often varies per tissue^64,65^. Our results, however, are derived from published variant to gene expression values (eQTLs). While we do see replication of a fraction of genes, when we compare the genes identified from our source GWASs for MRI traits (based on proximity to variants), our TWAS showed an overall increase in the number of genes associated for every MRI-trait tested. For example, from Pirruccello et al., 19 genes were identified as proximal to significant variants from GWAS for LVEF, while 27 unique genes were significant in a tissue specific context in TWAS^27^. This trend continued for LVEDV, LVESV, and LMV from Khurshid et al^26^. When we compare our gene results to additional previous GWASs for these traits available within GWAS Catalog^66^ and genes published with phenotypes in NCBI^67^, we also note the majority of our gene results of all four MRI traits are novel, indicating that a large information gap exists at the gene level for these traits. In relation to proteins, to our knowledge this is the first study looking at the protein level based on quantitative cardiac traits, making all results novel.

For all-cause multi-ancestry GWAS, 209 of the total 250 genes and proteins identified in our TWAS are considered novel associations and are not found in previous HF GWAS within GWAS Catalog (41 known genes and proteins replicated). 29 proteins were associated with HF. This increase in overall significant genes and proteins further increases the power of our gene set enrichments and protein-protein interaction networks.

### 4.2. Relationships between intermediate cardiac measures and HF

The four MRI derived cardiac measures explored in this study are often collectively used to mark structural and functional changes in the heart, and are reliable indicators of HF risk and eventual diagnosis. Therefore the relationship between these measures as well as the overlap between each and HF at the gene, protein, and pathway level is of interest.

Between the MRI cardiac measures we do see a certain amount of overlap at the gene and protein level. Three genes, *FKBP7*, *RP11-171I2.3*, and *PRKRA* were significant in at least one tissue for each of the four MRI cardiac traits. Both *FKBP7* and *PRKRA* have been discussed in relation to cardiac traits before. *PRKRA* has been implicated in studies impacting heart structure, and while *FKBP7* has been less discussed, was linked to atrial fibrillation in one study^68–72^. It is worth noting that *FKBP7*, *PRKRA*, and third gene (*PLEKHA3,* significant with LVM, LVEDV, and LVESV), all lie within a region of chromosome 2 that also includes the gene *TTN*, which is well established heritable cause of dilated cardiomyopathy, a leading cause of heart failure^73–75^.

Of the four traits LVEF and LVESV had the most overall overlaps, with 30 different genes and one protein, SPON1 appearing for both traits. When we clustered the results, one PPI cluster for LVEF is dominated by genes previously implicated with dilated cardiomyopathy and heart failure risk (*HSPB7*, *FLNC*, *ALPK3*, *CLCNKA*) **(Figure 4A)**, as well as links to the brain via *WDR73*^76–82^. We also see a cluster composed of much of the same genes for LVESV, and overlapping enriched pathways for cardiac cell development (*ALPK3*), and transepithelial chloride transport (*CLCNKA*). Renal transport also appears as a significantly enriched pathway for LVEF as a result of *CLCNKA,* as well as renal filtration cell differentiation and nephron tubule as top pathway results. Renal impairment is common among HF patients and is documented to increase mortality risk^83,84^.

Within our multi-ancestry HF results there appears to be substantial overlap in the MRI derived cardiac measures. 30 genes that appeared significant in one of the MRI cardiac traits also appeared to be associated with all-cause HF. LVEF and LVESV had the largest number of overlapping genes with the disease respectively (18 genes for LVEF and 18 genes for LVESV), further supporting their use as HF indicators. AIDA was the top significant protein from blood plasma, significant with both the European American and African American pQTL reference panels, a gene previously discussed as part of inflammatory response that also promotes atherosclerosis and coronary artery disease^85^. NCF1, which replicated at the gene and protein level, also has ties to immune response and is a key regulator of reactive oxygen species^86,87^. Other top proteins APOH, TNXB, PCSK9, and RGMB have previously been associated with HF^16,88–90^. Overall, HF associated gene and protein results tend to enrich lipid and specifically cholesterol related pathways. Nephron tubule development also appears as a top pathway for GF, implicating the kidneys again.

### 4.3. Limitations and future directions

There exist some limitations and several avenues for further analyses with this study. First, this analysis only encompasses the genetic factors of HF and intermediate cardiac measures. Genetics play a significant role in the development of HF; however, it has been shown that environment and comorbidities play a large role in increasing risk as well^91,92^. We did not consider social factors or health records of the patients used in the source GWASs, and therefore may not have fully been able to characterize all potential sources of disease progression. Future studies attempting to characterize or stratify individual-level risk of developing HF would benefit from including these data modalities, in addition to those used in this study.

Additionally, we used a GWAS study with an all-case HF multi-ancestry cohort to represent HF in our study. While this is beneficial for increasing sample size and increasing power, this might also muddle distinct signals within known phenotypic subgroups of HF or specific ancestries given the complexity of the disease as earlier stated. Future studies may find improvement by stratifying HF by subgroups, such as diastolic vs systolic dysfunction, to achieve more informed results. Beyond our cohort limitations, we also acknowledge that the imputation of gene and protein expression is influenced by the ancestry and completeness of the reference eQTL and pQTL sets we used (GTEx v8 and ARIC). It is possible some signals were left out in our results because they were not available in these QTL sets due to lack of representation. These QTL reference sets were also not disease specific. The creation of disease specific models using disease-specific RNAseq and proteomic data may be an avenue for further improvement. Lastly, here we only considered imputed gene and protein data modalities, based on multi-omics data from well characterized reference populations. The use of additional modalities, such as RNAseq, protein abundance, or methylation information would provide stronger evidence for our conclusions.

## Data Availability

All reference data used are available online through GTEx and ARIC. All summary statistics are accessible through their original published studies.

https://ritchielab.org/publications/supplementary-data/psb-2025/hfmultiomics

## Acknowledgments

This work was supported through the following grants from the National Institutes of Health (NIH): AG066833, HL169458. We also thank Tess Cherlin from the University of Pennsylvania for her assistance with visualizations in this manuscript.

## Appendix

Supplemental figures and tables are available at: https://ritchielab.org/publications/supplementary-data/psb-2025/hfmultiomics

