## Supplemental Figures for "Connecting intermediate phenotypes to disease using multi-omics in heart failure"

#### Supplemental Figure 1: Genomic, Transcriptomic, and Proteomic Associations - Additional Phenotypes

##### MRI – LV Mass

**A**

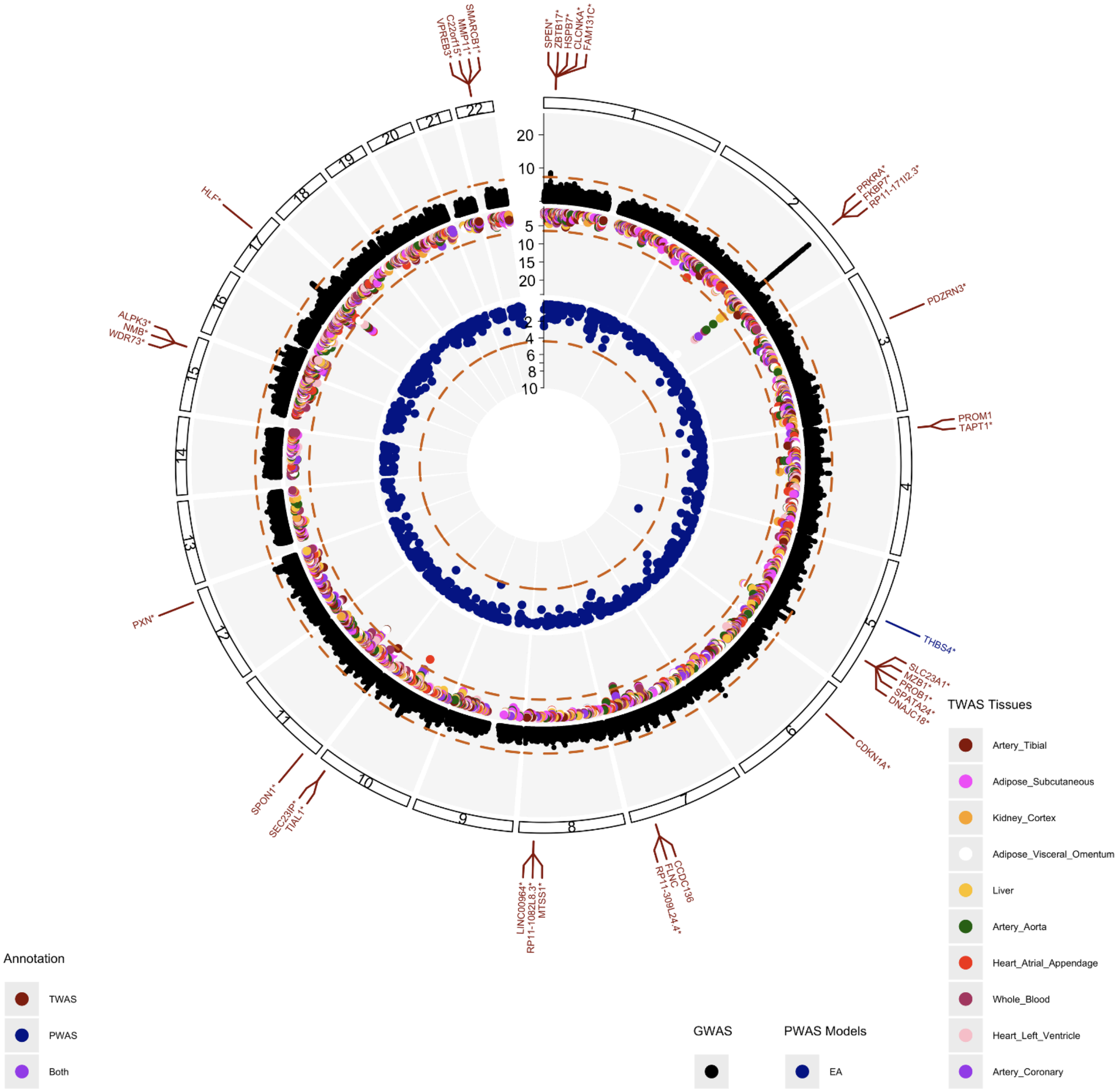

### MRI – LV End-Systolic Volume

**B**

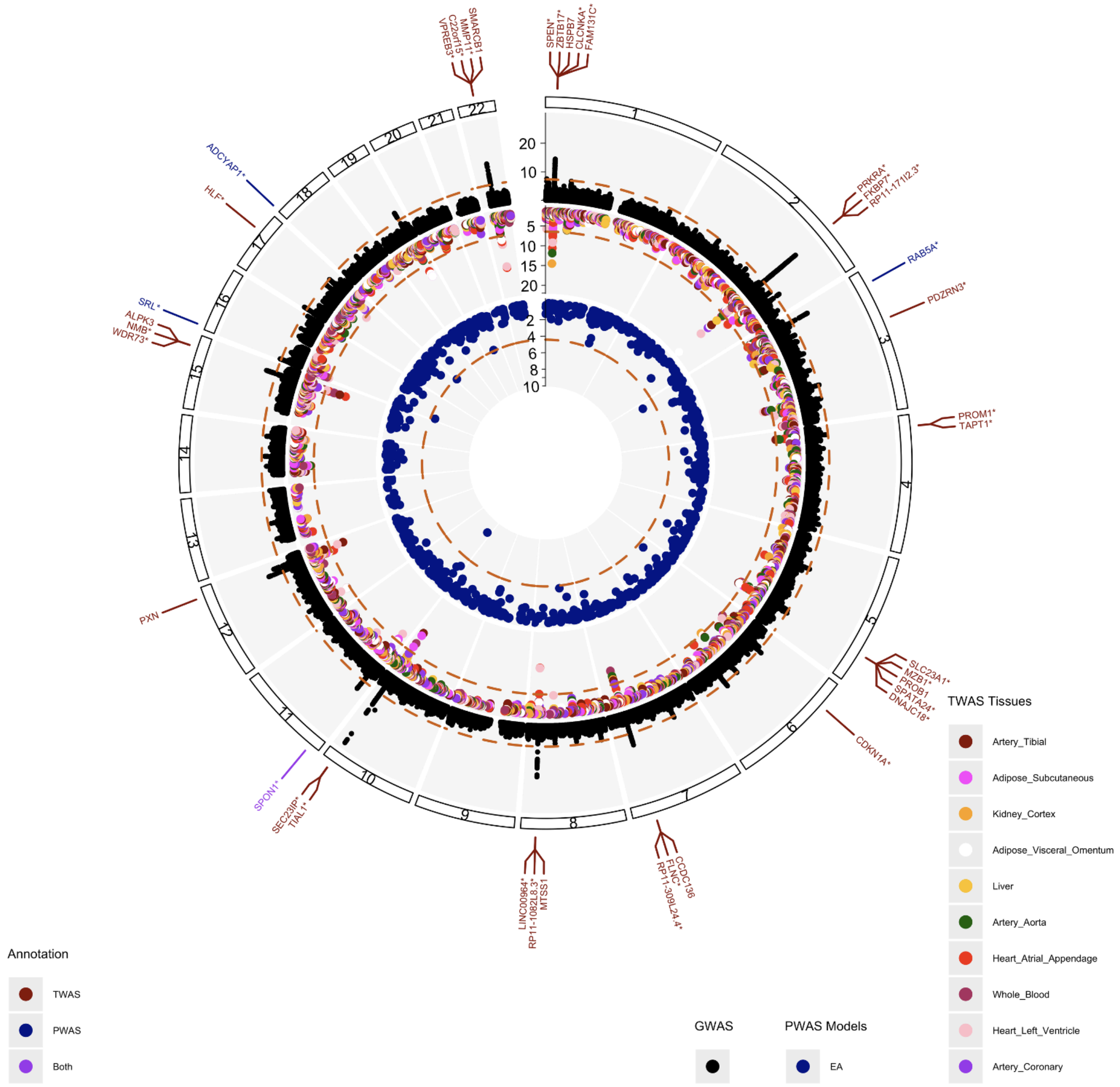

### MRI – LV End-Diastolic Volume

**C**

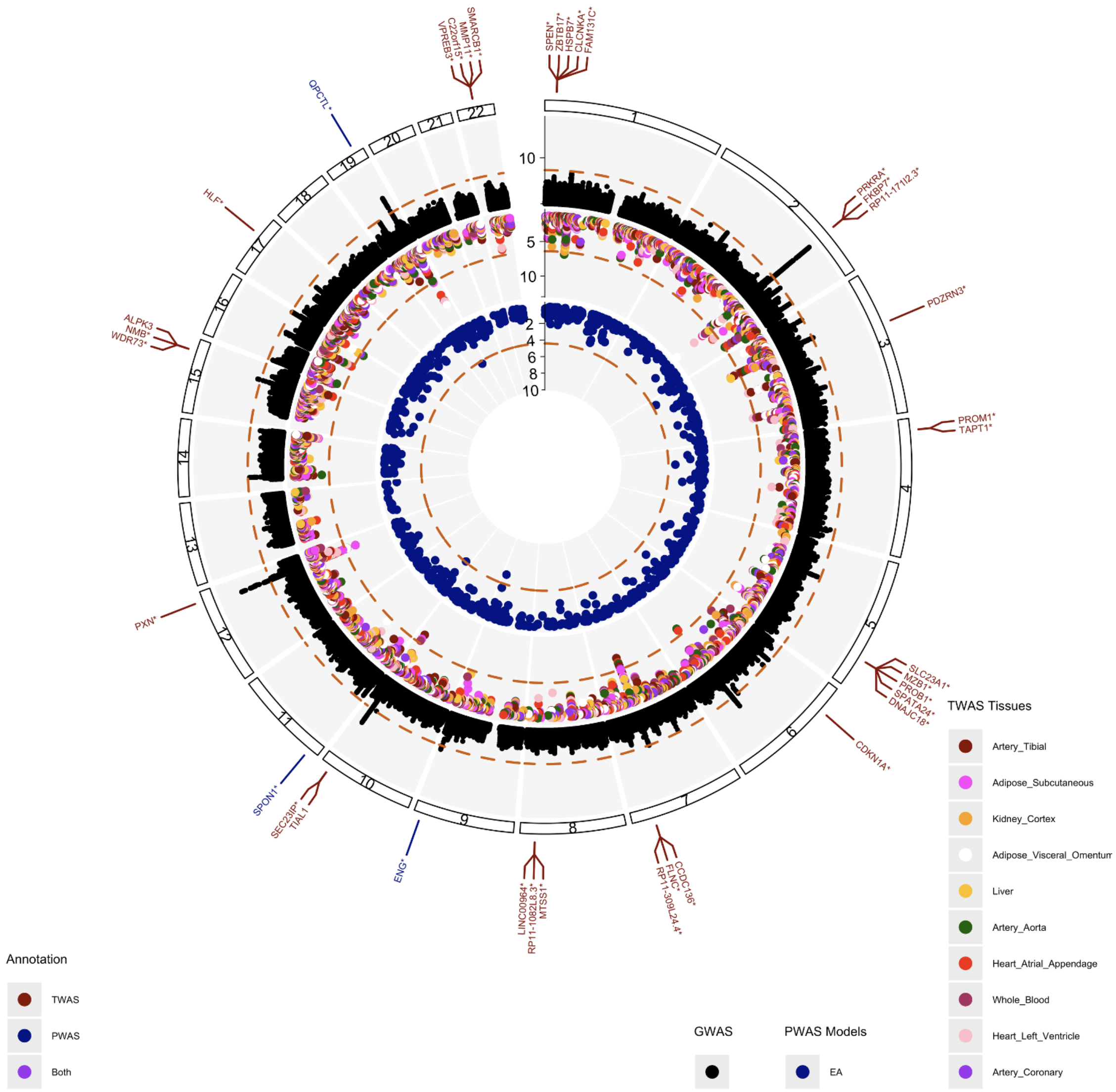

### HF - European

**D**

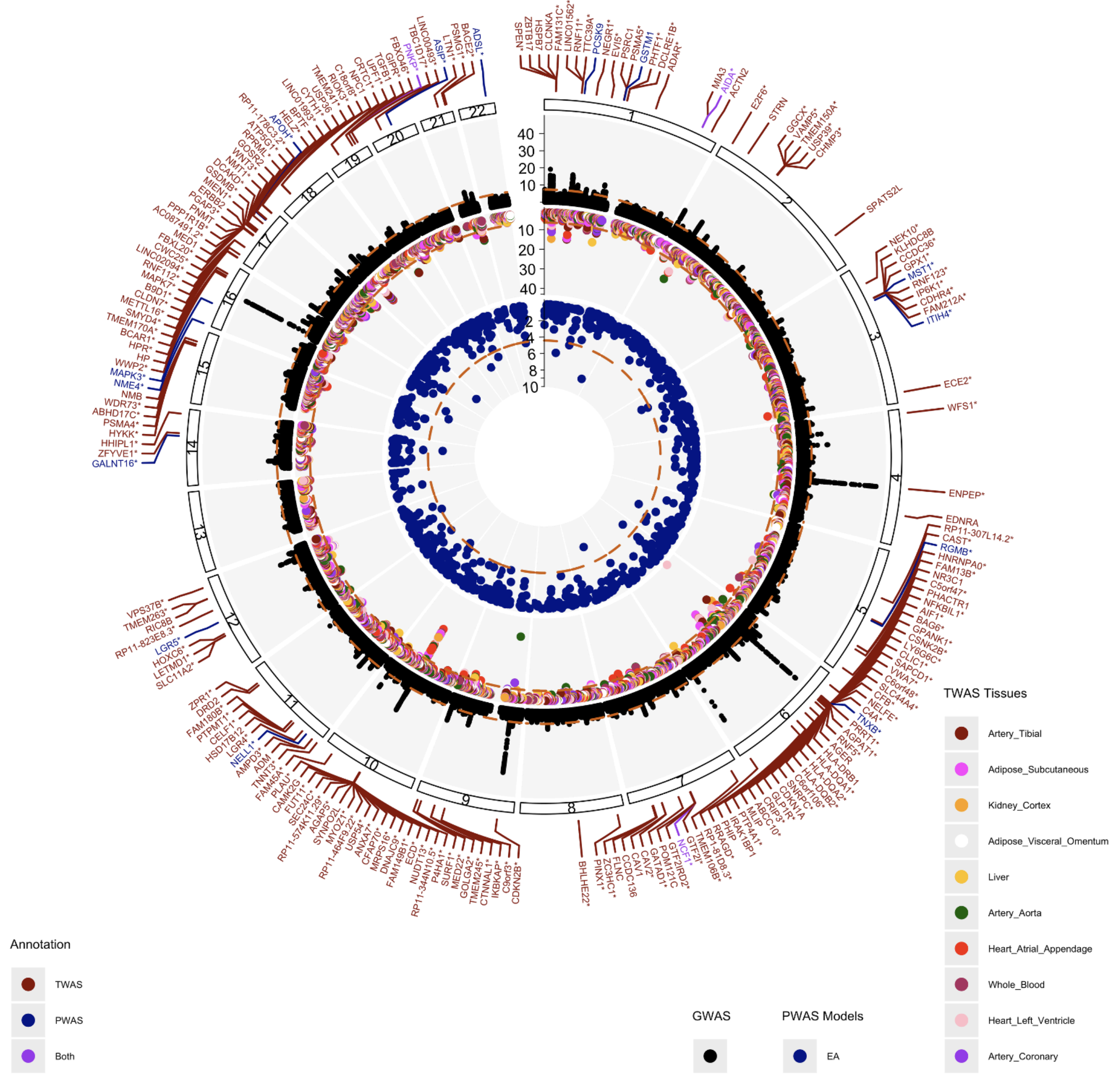


**Supplemental Figure 1: Circos plots for A) MRI LV Mass, B) MRI LV End-Systolic Volume, C) MRI LV End-Diastolic Volume, and D) HF - European representing identified associations through GWAS (black), TWAS (red), and PWAS (blue).**

#### Supplemental Figure 2: Gene-Set Enrichment Results - Additional Phenotypes

**A**

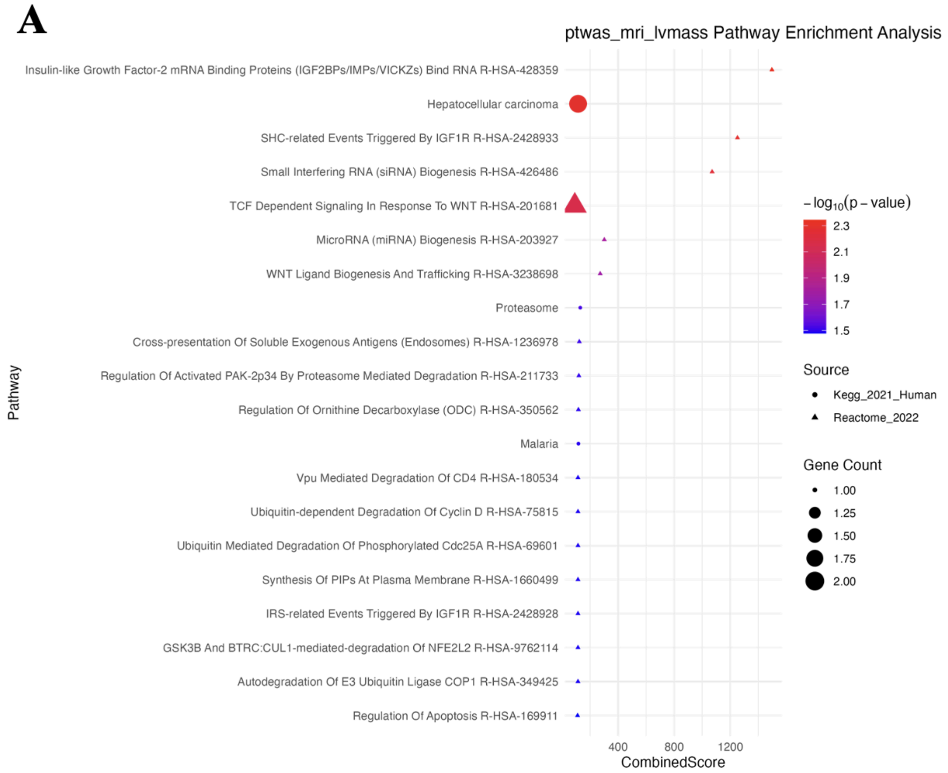


**B**

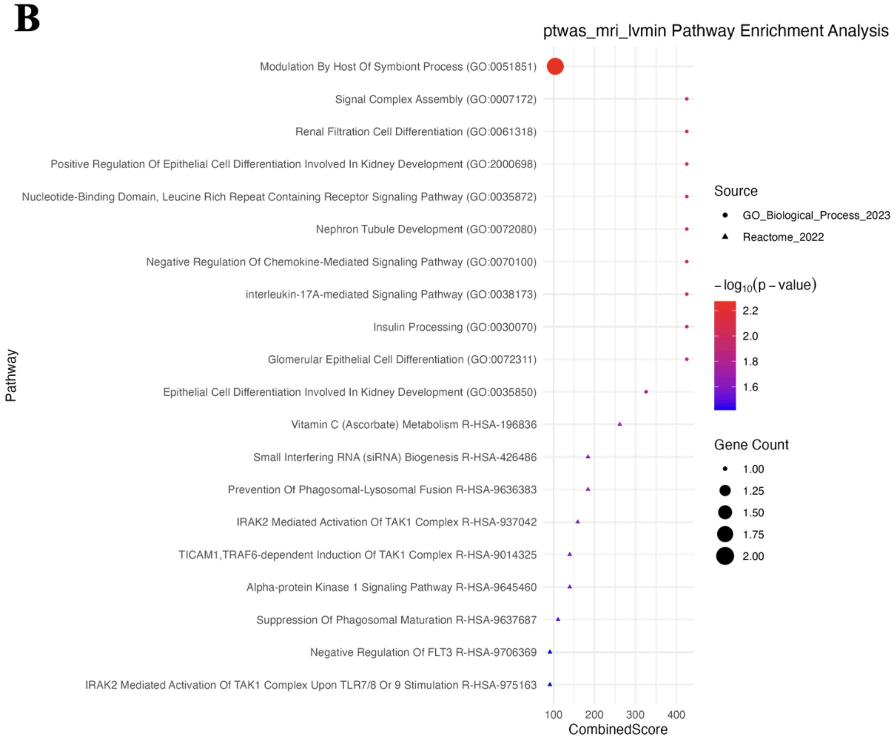

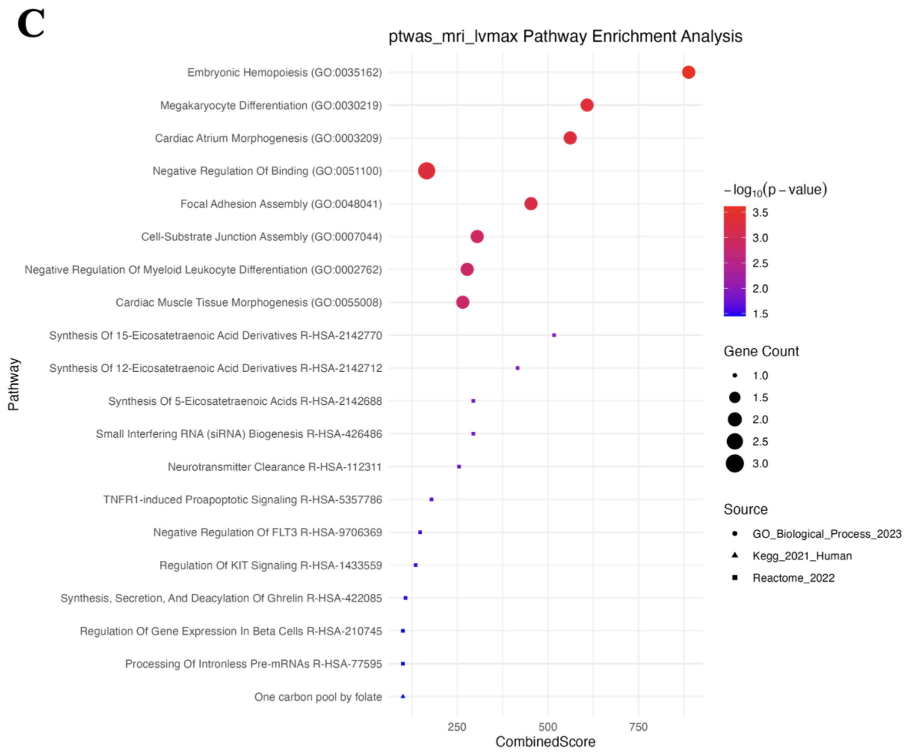


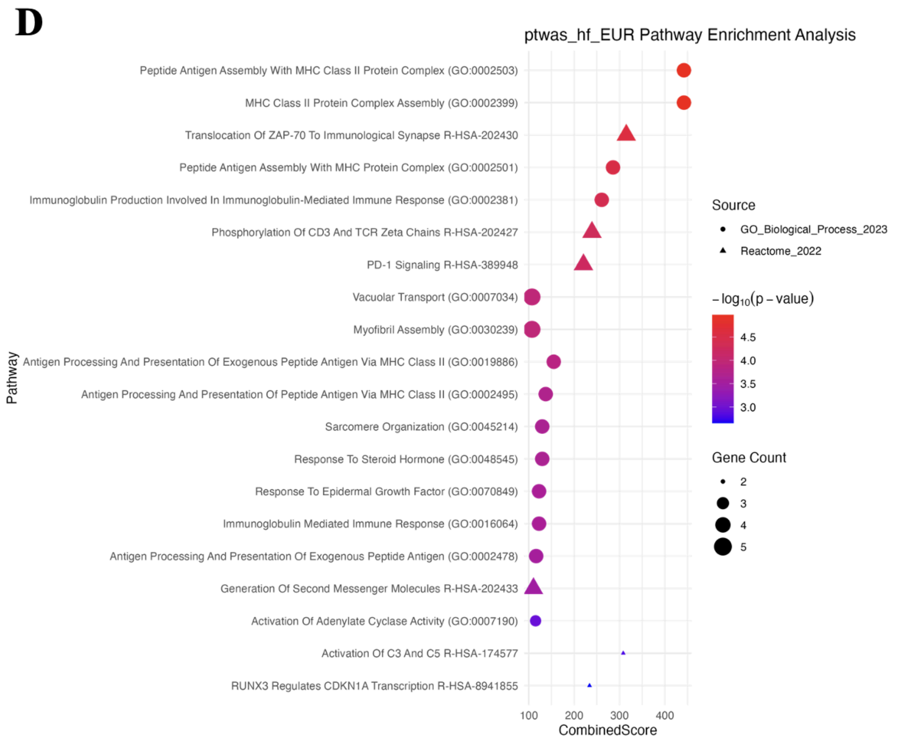


**Supplemental Figure 2: Gene-set enrichment results of TWAS and PWAS significant hits for A) MRI LV Mass, B) MRI LV End-Systolic Volume, C) MRI LV End-Diastolic Volume, and D) HF - European phenotypes.**

**Supplemental Figure 3: PPI Networks - Additional Phenotypes**

**MRI – LV Mass PPI Network**

**A**

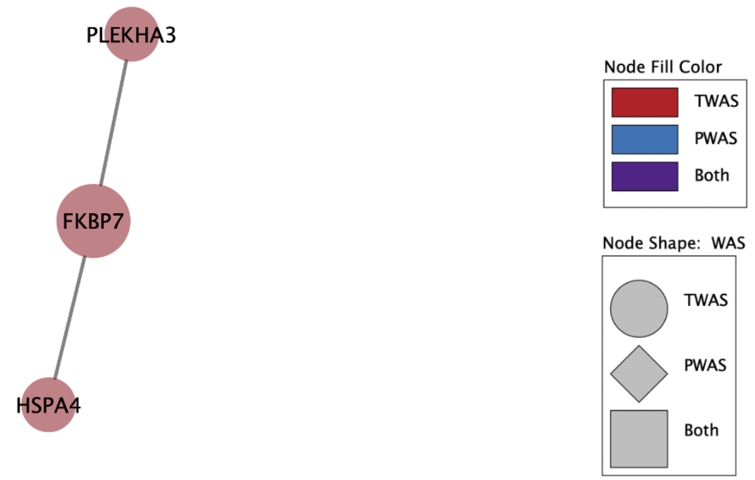


**MRI – LVESV PPI Network**

**B**

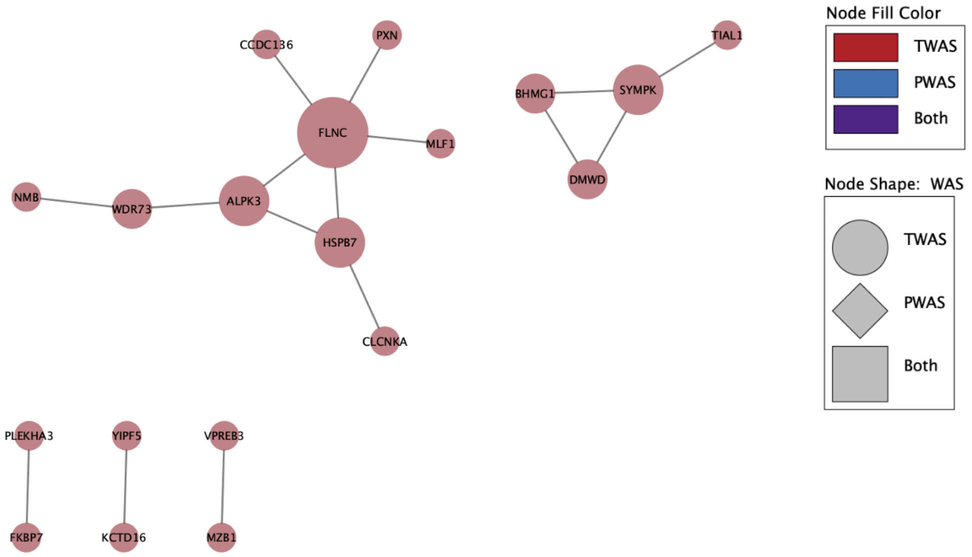

#### MRI – LVEDV PPI Network

**C**

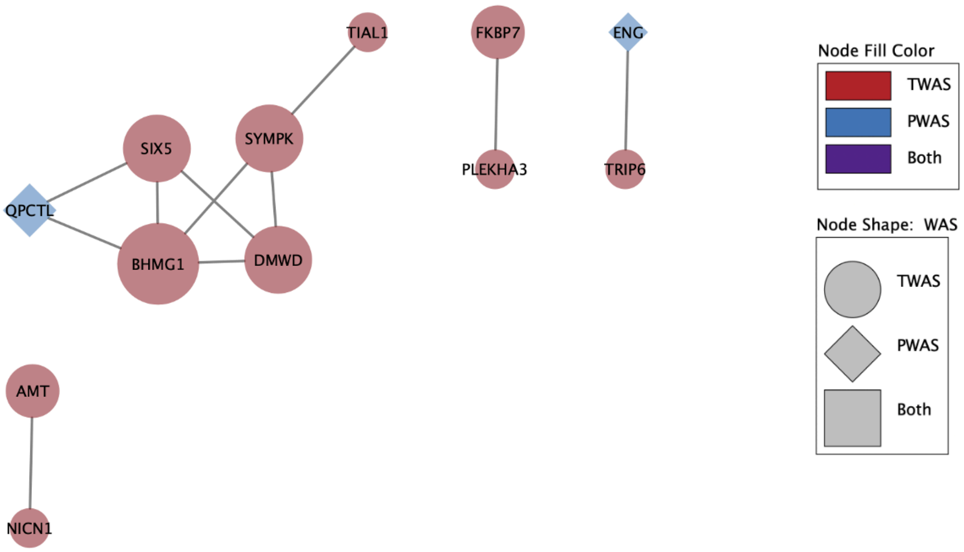


#### HF – EUR PPI Network

**D**

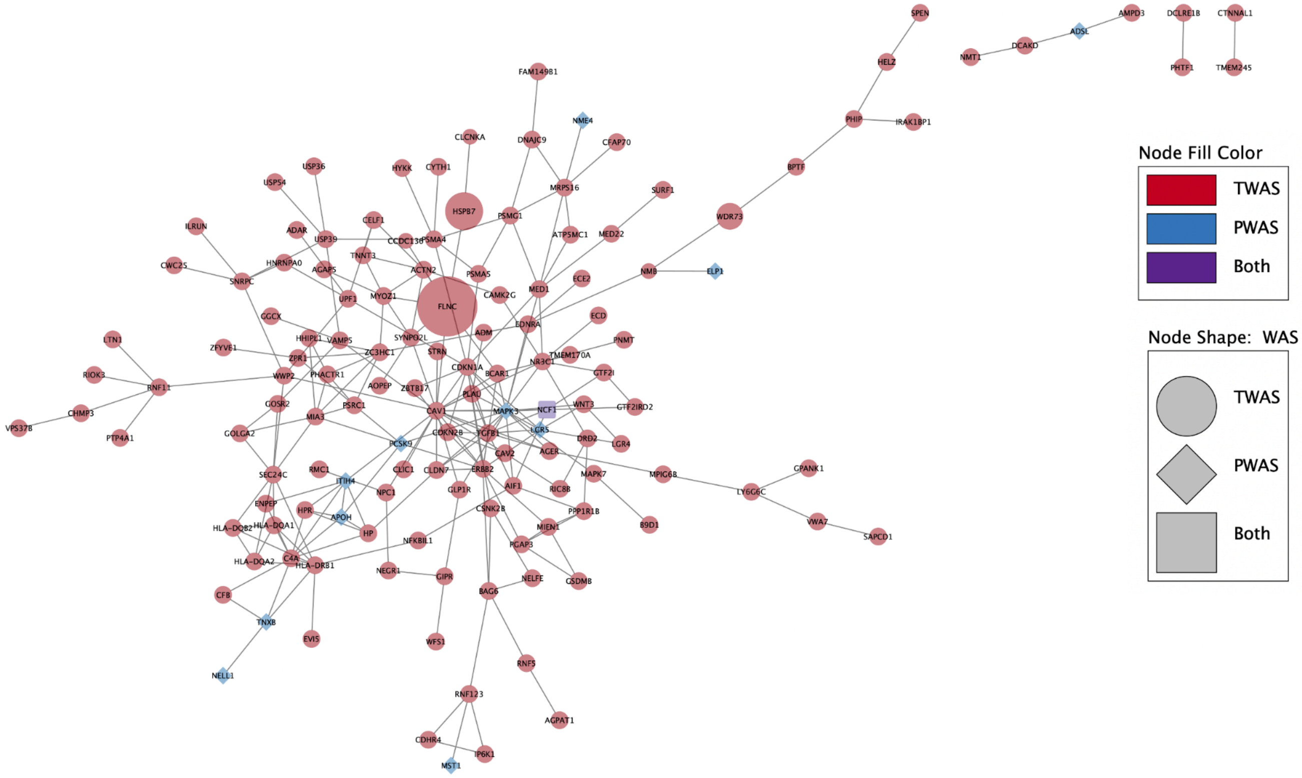


**Supplemental Figure 3: PPI networks constructed using TWAS and PWAS significant hits for A) MRI LV Mass, B) MRI LV End-Systolic Volume, C) MRI LV End-Diastolic Volume, and D) HF - European phenotypes**
